# Short Report –Consumption of multiple sources of protein is associated with gestational diabetes mellitus in the Chinese population

**DOI:** 10.1101/2021.07.27.21261201

**Authors:** Wei-Qin Wang, Vic Shao-Chih Chiang, Jing-Yuan Wen, Ji-Fen Hu, Rong-Xian Xu

## Abstract

**Background:** Gestational diabetes mellitus (GDM) is a hyperglycemic state during pregnancy that results in disruptions of insulin sensitivity and secretion. It affects 7% of all pregnancies and lead to adverse maternal and offspring outcomes. GDM has many risk factors, such as ethnicity special, hereditary. However Nutritional factors offer key strategies against GDM, but this is less characterized for the Chinese population.

**Methods:** A case control study of GDM pregnant women (n=49, 29.88±3.92 years of age) and healthy pregnant women (n=77, 27.63±4.83 years of age) from 1^st^ Affiliated Hospital of Fujian Medical University were studied. Diagnosis was made using oral glucose tolerance test. Dietary data were collected using food frequency questionnaires. Data for use of dietary supplements, occupation, education, income, meal expenditure and smoking history were also recorded.

**Results:** No differences were found between GDM and control subjects for their age, education, occupation, monthly income, grocery expenditure and smoking (p>0.05). GDM subjects were associated with higher quantities of dairy products (p<0.05) and seafood (p<0.01) intake. There were also higher number of GDM subjects using protein powder supplementation (p<0.05).

**Conclusions:** Dietary recommendation guidelines for pregnant Chinese women should consider possible risks with excessive consumption of dairy products, seafood and protein powder. They should also assess the quality of the nutrition factor, other dietary interactions and their nutritional status further to minimize adverse outcomes caused by GDM.

## Introduction

Gestational diabetes mellitus (GDM) is a state of hyperglycemia with onset or first identification during pregnancy (Farrar, *et al*., 2015). It affects over 7% of all pregnancies and increases risks for maternal and perinatal mortality or health consequences later in life (Farrar, *et al*., 2015).

This state of hyperglycemia presents symptoms of polyuria, polydipsia, polyphagia, weight loss and blurred vision (American Diabetes Association, 2014). Its diagnosis is evaluated based on glucose tests and its criteria threshold has been reviewed many times (Meek, *et al*., 2015). GDM attributes to multifactorial pathophysiologies involving β-cell function, insulin signaling, pregnancy hormones and growth factors that result in disruptions of insulin sensitivity and secretion (Lappas, *et al*., 2011). Several factors are known to increase risks of GDM such as family history of type 2 diabetes mellitus, maternal age and maternal body mass index (Teh, *et al*., 2011). Well-established strategies against GDM include physical exercise, insulin therapy and oral medicating metformin (Artal, 2015,Zhao, *et al*., 2015).

Nutritional factors offer key strategies to combat GDM including probiotics, *myo-* inositol, fibre, reduced energy intake and reduced glycemic load (Rogozinska, *et al*., 2015). However, there are limited amount of research surrounding this topic in the Chinese population. This is important because different ethnicities present disparities in GDM risks and outcomes (Chakkalakal, *et al*., 2015). Dietary recommendations should therefore be customized for different ethnicities in order to optimize improve in the prevalence of GDM. This case-control study aimed to investigate food intake and dietary supplementation in Chinese GDM subjects.

## Methods

### Overview

The case control study included pregnant women that received prenatal physical examinations and awaits delivery at the 1^st^ Affiliated Hospital of Fujian Medical University. The exclusive criteria were listed as follows: recent respiratory and urinary tract infections, external injuries, hemorrhage, other stress conditions, presence of tumors, transmissible diseases (e.g. tuberculosis, hepatitis B) and chronic diseases (e.g. connective tissue diseases). The subjects were divided into GDM group (n=49) and the control group (n=77) in a 1 to 1.5 ratio mismatched design. The study was approved by the Biomedical research Ethics Committee at Fujian Medical University ([2011] Fu Zi Medical Ethics Review (52)).

### Gestational Diabetes Mellitus Diagnosis

The diagnosis of GDM was determined using 75g oral glucose intolerance test (OGTT) at fasting and post-challenge (one and two hours). GDM was identified if any of the OGTT measurements exceeded the specified threshold. These were: 5.1mmol/L (fasted), 10.0 mmol/L (one hour post-challenge) and 8.5 mmol/L (two hour post-challenge). These tests were conducted at gestational stage of approximately 24 weeks.

### Collection of Dietary Information

The dietary habits of the subjects were investigated using food frequency questionnaires. The questionnaire was origined with guidance from the 2010 Chinese citizen nutrition and health status investigations for pregnant women. The questionnaire was composed of 11 major food types: staple foods, beans, vegetables, algae, fruits, dairy, meat, seafood, egg, snacks and beverages. Records were made for the frequency and quantity of consumption. The uses of dietary supplements were also recorded. Further records were taken for possible confounders: age, occupation, education, income, meal expenditure and smoking history.

### Statistical Analysis

The data were statistically analyzed using SPSS statistics 17 software. Using t-tests, the differences in the consumption of various food types and dietary supplements were compared. Chi square tests were performed for comparisons of the potential confounding variables.

## Results

There were no differences between GDM and control subjects in potential confounding variables (p>0.05): age, location, education, occupation, monthly income, grocery expenditure and smoking (Table 1). In general, the recruited subjects were 29 years of age, living in city area and received secondary or undergraduate education. In terms of occupation, most were company clerks or stays at home, with income generally over 6000 CNY per month. In terms of meal expenditure, 10 – 30% of income for most of them goes towards this expense. The recruited subjects were overall non-smokers.

**Table 1.** Social background of recruited subjects

|  | GDM (n=49) | Control (n=77) | P-value |
| --- | --- | --- | --- |
| Age | 29.88 ± 3.92 | 27.63 ± 4.83 | 0.264 |
| Location |  |  | 0.825 |
| Sea Coast | 2(1.6%) | 3(2.4%) |  |
| Rural | 9(7.1%) | 11(8.7%) |  |
| Urban | 38(30.2%) | 63(50.0%) |  |
| Education |  |  | 0.989 |
| Elementary | 1(0.8%) | 2(1.6%) |  |
| Intermediate | 9(7.1%) | 12(9.5%) |  |
| Secondary | 11(8.7%) | 18(14.3%) |  |
| Undergraduate | 25(19.8%) | 39(31.0%) |  |
| Postgraduate | 3(2.4%) | 6(4.8%) |  |
| Occupation |  |  | 0.888 |
| Civil Service | 1(0.8%) | 2(1.6%) |  |
| Company Clerk | 14(11.1%) | 22(17.5%) |  |
| Professionals | 9(7.1%) | 16(12.7%) |  |
| Worker/ Service | 1(0.8%) | 0(0%) |  |
| At-Home | 18(14.3%) | 28(22.2%) |  |
| Other | 6(4.8%) | 9(7.1%) |  |
| Monthly Income |  |  | 0.816 |
| ≤ 3000 CNY | 1(0.8%) | 2(1.6%) |  |
| 3000 - 6000 | 11(8.7%) | 15(11.9%) |  |
| ≥6000 CNY | 36(29.4%) | 60(47.6%) |  |
| Meal Expenditure (Ratio to Income) |  |  | 0.191 |
| < 10% | 7(5.6%) | 4(2.4%) |  |
| 10 - 30% | 35(28.6%) | 61(48.4%) |  |
| 30 - 50% | 5(4.0%) | 11(8.7%) |  |
| > 50% | 1(0.8%) | 2(1.6%) |  |
| Smoking History |  |  | 0.126 |
| Yes | 1(0.8%) | 4(3.2%) |  |
| No | 48(38.1%) | 73(57.9%) |  |
| Secondhand Smoke |  |  | 0.124 |
| Yes | 4(3.2%) | 15(11.9%) |  |
| No | 45(35.7%) | 62(49.2%) |  |

The average daily intake of different food types between GDM and control subjects were recorded (Table 2). In terms of dairy consumption, GDM subjects (301.37± 235.46) had higher average daily consumption than control subjects (188.72±168.25) (p<0.05). Higher consumption were also observed for seafood in GDM subjects (158.03±168.36) compared to control subjects (100.25±60.51) (p<0.01). No differences were found for other food types that were recorded (p>0.05).

**Table 2.** Average daily intake of different food types of recruited subjects

| Food Type | GDM (n=49) | Control (n=77) | P-value |
| --- | --- | --- | --- |
| Staple food (g) | 356.95±110.62 | 367.40±94.66 | 0.450 |
| Beans (g) | 25.10±19.42 | 29.78±38.37 | 0.253 |
| Vegetables (g) | 254.65±140.44 | 275.14±148.34 | 0.513 |
| Algae (g) | 35.22±24.67 | 28.29±23.18 | 0.254 |
| Fruit (g) | 493.31±260.01 | 628.40±292.20 | 0.581 |
| Dairy (g) | 301.37±235.46 | 188.72±168.25 | 0.032 * |
| Meat (g) | 132.47±96.19 | 102.70±74.40 | 0.167 |
| Seafood (g) | 158.03±168.36 | 100.25±60.51 | 0.007 ** |
| Egg (g) | 52.90±29.74 | 45.97±26.26 | 0.566 |
| Snacks (g) | 88.63±70.18 | 97.72±80.06 | 0.684 |
| Beverages (g) | 8.70±19.07 | 20.01±79.91 | 0.135 |

Statistical significance of difference between GDM and control \* p<0.05; \*\* p<0.01

The use of different dietary supplements in both GDM and control subjects were recorded (Table 3). Supplementation with protein powder was associated with GDM (p<0.05). Other dietary supplements that were utilized were not different between GDM and control subjects (p>0.05).

**Table 3.** Use of different dietary supplements by recruited subjects

|  | GDM (n=49) | Control (n=77) | P-value |
| --- | --- | --- | --- |
| Folic acid |  |  | 0.483 |
| Yes | 32(27.1%) | 61(51.7%) |  |
| No | 11(9.3%) | 14(11.9%) |  |
| Iron |  |  | 0.094 |
| Yes | 8(6.8%) | 25(21.2%) |  |
| No | 35(29.7%) | 50(42.4%) |  |
| Calcium |  |  | 0.287 |
| Yes | 34(28.8%) | 66(55.9%) |  |
| No | 9(7.6%) | 9(7.6%) |  |
| Multivitamin |  |  | 1.000 |
| Yes | 24(20.3%) | 43(36.4%) |  |
| No | 19(16.1%) | 32(27.1%) |  |
| Formula milk |  |  | 0.413 |
| Yes | 8(6.7%) | 12(10.2%) |  |
| No | 35(29.7%) | 63(53.4%) |  |
| Protein powder |  |  | 0.011* |
| Yes | 7(5.9%) | 2(1.7%) |  |
| No | 36(30.5%) | 73(61.9%) |  |
| Bird's nest |  |  | 0.157 |
| Yes | 0(0%) | 5(4.2%) |  |
| No | 43(36.4%) | 70(59.3%) |  |
| Honey |  |  | 0.364 |
| Yes | 1(0.8%) | 0(0%) |  |
| No | 42(35.6%) | 75(63.6%) |  |

Statistical significance of difference between GDM and control \* p<0.05

## Discussion

Recommendations for GDM need to be ethnicity-specific due to distinct risks (Chakkalakal, *et al*., 2015). Nutrition is an imperative strategy to combat GDM (Rogozinska, *et al*., 2015) and this case-control study aimed to investigate food intake and dietary supplementation in Chinese GDM subjects.

The association of dairy with GDM found in the present study is novel. However, it contradicts current recommendations to increase dairy intake for GDM prevention (Rono, *et al*., 2014). Furthermore, a systematic review found dairy to benefit type 2 diabetes (T2M), which has similar pathophysiology to GDM (Turner, *et al*., 2015). It is important to note that the dairy in these studies were low-fat. Since high fat diet increases risks of GDM (Park, *et al*., 2013), the incorporation of all types of dairy in the present study may have resulted in this contradiction.

Mediterranean diet is known to improve outcomes of GDM and it works in part through increasing seafood intake (Perez-Ferre, *et al*., 2014). Data on seafood association with GDM is non-existent but the present study failed to concord with this rationale. This is may be due to the major issue of seafood contamination by toxic metals in China (Du, *et al*., 2012). Some of these toxic metals have been previously associated with GDM (Romano, *et al*., 2015), but these were not accounted for in the present study.

Low protein intake increases risk of GDM (Ignacio-Souza, *et al*., 2013). Protein powder strategizes to solve this problem and has been found to improve T2M (Jakubowicz, *et al*., 2014). In contrast, the present study found supplementation of protein powder to associate positively with GDM. This divergence may stem from the non-quantitative assessment of dietary supplement use. The dose is critically important as excessive supplementation in addition to whole food proteins may actually be counterproductive (Rehfeldt, *et al*., 2014).

It is interesting to note that the nutritional associations found in the current study were all considered as major sources of protein (Bao, *et al*., 2013). High protein diets have been systematically reviewed to benefit T2M (Dong, *et al*., 2013). In saying that, this could not be further probed in this study since nutrient-based approaches were not adopted. This notion is complicated by its dependency on the type of protein because animal protein demonstrates greater GDM risk to vegetable proteins (Bao, *et al*., 2013). Caveats should likewise be taken that substantial ancillary protein intake in protein-sufficient subjects would *de facto* be detrimental.

Limitations of this study include its sample size of GDM cases and the above mentioned characterization of nutritional items. Nonetheless, it addresses important nutritional considerations to oppose GDM in the limitedly investigated Chinese population, to offer grounding for future research. The nutritional items investigated should be more defined in future research includes: low fat products, metal contamination, dietary supplementation dose and overall macronutrient intake. The nutritional status of the subjects should be similarly considered.

## Conclusion

Since risks of GDM are ethnicity-dependent (Chakkalakal, *et al*., 2015), dietary recommendations need to be more tailored to different ethnicities. The consumption of dairy, seafood and protein powder needs attention for pregnant Chinese women. Considerations need to be made for the nutritional status, fat content, metal contamination, quantity and balance with other dietary intakes. GDM affects the l pregnancies (Farrar, *et al*., 2015) and diet offers crucial GDM strategies to reduce the subsequent GDM health complications and mortalities (Rogozinska, *et al*., 2015).

## Data Availability

The datasets generated during and/or analysed during the current study are available from the corresponding author on reasonable request.

## Acknowledgements

The authors would like to thank the participation of pregnant women and the assistance of the nurses at the obstetric ward.

## Conflict of interests, sources of funding and authorship

The authors declare that they have no competing interests.

This study was supported by Fujian Province Natural Science Fund 2012-2014.

Rong-xian Xu was in charge of the study design and funding applications. Wei-Qin Wang conducted the study, facilitated data collection, data analyses and report-writing. Vic Shao-Chih Chiang and Jing-Yuan Wen wrote and reviewed the manuscripts. Ji-Fen Hu managed the nurses for subject recruitments.

